# *DPYD* genetic polymorphisms in non-European patients with severe fluoropyrimidine-related toxicity: A systematic review

**DOI:** 10.1101/2023.12.11.23299813

**Authors:** Tsun Ho Chan, J. Eunice Zhang, Munir Pirmohamed

## Abstract

**Background:** Pre-treatment *DPYD* screening is mandated in the UK and EU to reduce the risk of severe and potentially fatal fluoropyrimidine-related toxicity. Four *DPYD* gene variants which are more prominently found in Europeans are tested.

**Methods:** Our systematic review in patients of non-European ancestry followed PRISMA guidelines to identify relevant articles up to April 2023. Published *in silico* functional predictions and *in vitro* functional data were also extracted. We also undertook *in silico* prediction for all *DPYD* variants identified.

**Results:** In 32 studies, published between 1998 and 2022, 53 *DPYD* variants were evaluated in patients from 12 countries encompassing 5 ethnic groups: African American, East Asian, Latin American, Middle Eastern, and South Asian. One of the 4 common European *DPYD* variants, c.1905+1G>A, is also present in South Asian, East Asian and Middle Eastern patients with severe fluoropyrimidine-related toxicity. There seems to be relatively strong evidence for the c.557A>G variant, which is found in individuals of African ancestry, but is not currently included in the UK genotyping panel.

**Conclusion:** Extending UK pre-treatment *DPYD* screening to include variants that are present in some non-European ancestry groups will improve patient safety and reduce race and health inequalities in ethnically diverse societies.

## Introduction

Fluoropyrimidines are antimetabolite chemotherapy drugs comprising the parenterally administered 5-fluorouracil (5-FU) and its prodrugs, capecitabine and tegafur. They are commonly used either as monotherapy or in combination with other antineoplastic agents in neo-adjuvant, adjuvant and palliative settings for a variety of solid tumour types including colorectal, breast, oesophago-gastric and head and neck cancers ^1,2^. 5-FU and capecitabine have been on the World Health Organization (WHO) Essential Medicines List (EML) since 1977 and 2015, respectively ^3,4^. Annually, over two million patients worldwide and approximately 600,000 patients in Europe receive treatment with fluoropyrimidines ^5–7^. Due to a narrow therapeutic index, 10-30% of patients who receive standard fluoropyrimidine doses develop severe toxicity including bone marrow suppression, diarrhoea, mucositis and hand-foot syndrome, usually within the first 1-2 cycles of treatment ^8–11^. Severe fluoropyrimidine-related toxicity leads to mortality in approximately 0.5-1% of patients (with up to 5% lethal toxicity reported in elderly patients) ^12–16^.

Development of toxicity is in part due to inter-individual variability in dihydropyrimidine dehydrogenase (DPD) activity. The first case report of a patient presenting with 5-FU-related severe toxicity due to DPD deficiency was in 1985^17^. DPD is the primary enzyme responsible for the catabolism and elimination of >80% of the administered 5-FU to the inactive metabolite dihydrofluorouracil (DHFU)^1,15,18,19^. Deficiency of the DPD enzyme, either complete or partial, leads to inadequate clearance of 5-FU which increases drug exposure and accumulation, increasing the risk of severe and sometimes fatal toxicity ^20–22^. DPD deficiency can be detected in 39–61% of patients with severe fluoropyrimidine-related toxicity ^23^. In individuals of European ancestry, the frequency of partial DPD enzyme deficiency ranges from 3 to 5% while complete DPD enzyme deficiency is less frequent, with an estimated prevalence of 0.1-0.2% ^24,25^.

The DPD gene (*DPYD*) is expressed in a wide variety of human tissues; high levels are observed in the liver and peripheral blood mononuclear cells (PBMCs) ^26,27^. Located on chromosome 1p21.3, *DPYD* is a large pharmacogene spanning ∼920 kb in length, with 23 relatively small exons (69-961 bp) surrounded by large intronic regions ^28,29^. The coding sequence totals ∼3 kb in length and encodes a polypeptide comprising 1,025 amino acid residues ^28,29^. *DPYD* is highly polymorphic: the Genome Aggregation Database (gnomAD v2.1.1) includes 204 synonymous variants and 569 missense variants, 40 of which are predicted to lead to loss of enzymatic function ^30^.

The latest version of the Clinical Pharmacogenetics Implementation Consortium (CPIC) guideline includes 82 known *DPYD* variants, among which, 21 are considered to have no DPD function and 6 to have diminished DPD function ^6^. Prospective genotyping of *DPYD* can identify patients with DPD enzyme deficiency and allow for prophylactic fluoropyrimidine dose adjustments, thereby reducing the likelihood of fluoropyrimidine-related toxicity without compromising cancer treatment effect ^31–35^.

In June 2020, the European Medicines Agency (EMA) recommended DPD testing either by phenotyping or genotyping prior to treatment with fluoropyrimidines ^36^. In November 2020, the National Health Service (NHS) commissioned *DPYD* genetic testing making this one of the first pharmacogenomic tests to be applied nationally in the UK ^37^. A variety of genotyping methods are used by the labs but they all test for the four pathological *DPYD* variants commonly described in Europeans:

- c.1905+1G>A (IVS14+1G>A, rs3918290, *DPYD* *2A), a splice-site variant causing exon 14 skipping which results in the production of an inactive protein ^38,39^;
- c.2846A>T (p.Asp949Val, rs67376798, *DPYD*\*9B), a non-synonymous variant that leads to reduced DPD activity;
- c.1236G>A/HapB3 (p.Glu412=, rs56038477), a synonymous variant which tags for c.1129-5923C>G (rs75017182), a deep-intronic splice-site variant causing significant loss of DPD activity, which is in near perfect linkage disequilibrium (LD) with the *DPYD* haplotype HapB3 encompassing three intronic variants (rs56276561, rs6668296, rs115349832); and
- c.1679T>G (p.Ile560Ser, rs55886062, *DPYD*\*13), a missense variant causing decreased DPD activity.

This is because the three key clinical studies which provided evidence for the clinical utility of *DPYD* testing to reduce the incidence of severe fluoropyrimidine-related toxicity were all undertaken in European populations ^11,31,32^. The minor allele frequencies (MAF) of these four prominent European *DPYD* variants across non-European population groups from the 1000 Genomes Project Phase 3 ^40^ and gnomAD v3.1.2 and v4.0.0 ^41^ databases are shown in Supplementary Table 1.

It is known that there are inter-ethnic differences in *DPYD* variant frequency. In fact, several studies have reported the absence of the European *DPYD* variants in populations from East and Southern Africa, namely Somalia, Kenya ^42^ and Zimbabwe ^43^, and East Asia including China ^44^ and Japan ^45–48^. In addition, variants that are not present in Europeans can have a profound impact in non-European populations, and vice versa ^49^. Hence, the testing being undertaken by EU countries and the UK NHS will not identify genetic variants in non-European populations, who will be treated as wild-type, and given conventional doses of the fluoropyrimidine drugs, with the likelihood of toxicity, and in the worst-case scenario, death. This has the potential to exacerbate health and race inequalities in ethnically diverse societies. Furthermore, it does not help countries where the population is predominantly of non-European ancestry, as *DPYD* genetic testing will not be implemented because of lack of evidence. It is crucial that all global populations benefit equally from this important genetic test. We have therefore undertaken a systematic review to evaluate *DPYD* genetic variants which have been reported in patients of non-European ancestry who developed severe fluoropyrimidine-related toxicity.

## Methods

### Design and registration

A systematic review was conducted in accordance to the Preferred Reporting Items for Systematic reviews and Meta-Analyses (PRISMA) 2020 guideline ^50^. The review protocol was registered in the PROSPERO repository of systematic reviews (registration number CRD42023385227). The EndNote™ X9 software was used to manage all articles (both included and excluded records) throughout the research process.

### Search strategy

A literature search was performed using the MEDLINE (PubMed), Web of Science, Embase (OVID) and Scopus electronic databases to identify relevant articles published prior to 04 April 2023. The search strategy employed a combination of MeSH terms and keywords using the Boolean operators “AND” and “OR”. In addition, syntax adjustments were made appropriate to each database. The search terms used in the MEDLINE (PubMed) search are described in Supplementary Table 2; similar terms were used in the Web of Science, Embase (OVID), and Scopus searches.

### Eligibility criteria

We limited our search to clinical research studies, case series and case reports that genotyped for *DPYD* genetic variants in patients of non-European ancestry who had developed severe fluoropyrimidine-related toxicity after chemotherapy treatment containing 5-FU, capecitabine or tegafur. We accepted the definition of severe toxicities as (1) grade ≥3 severe adverse events according to the Common Terminology Criteria for Adverse Event (CTCAE) ^51^, (2) grade ≥3 severe adverse events in accordance to the World Health Organization (WHO) ^52^, and (3) dose-limiting toxicity (DLT) which is defined as pre-specified severe adverse events of grade 3 or higher based on the CTCAE classification. In order to maximise the number of included studies, we also accepted author-defined severity grading of fluoropyrimidine-related toxicities where terms ‘grade 3’, ‘grade ≥3’, ‘grade 4’, or ‘severe’ were used but no classification tool was specified.

Only publications with full text availability were included. Publications in all languages were assessed with non-English articles translated either via Google Translate or with assistance from colleagues who were native speakers of the foreign language. Authors and titles of conference meeting abstracts were used to check whether full-text articles had been published. Editorials, opinion letters, and unrefereed preprints were not considered.

### Screening process and study selection

After study duplications were removed, T.H.C screened the titles and abstracts of all articles in accordance with the above eligibility criteria to identify the relevant studies for first phase inclusion; irrelevant studies were excluded. In the second phase of the review process, full-text articles of the relevant studies were retrieved, and in-depth full-text screening was carried out. Detailed full-text screening also included the inspection of all cited references. In addition, the reference lists of clinical guidelines, policy statements from regulatory agencies, pertinent narrative and systematic reviews were also screened to check for additional eligible studies. In the situation of any uncertainty during the selection process, the full text was checked and resolved by consensus with J.E.Z.

### Quality assessment

T.H.C and J.E.Z independently assessed the methodological quality of each included study and relied on peer-review to ensure included studies were methodologically sound. A formal assessment of the risk of bias was not undertaken.

### Data extraction

Relevant summary and patient-level data from published manuscripts and appendix materials of included studies were independently extracted by T.H.C and J.E.Z. A data extraction form was compiled and data items collected are detailed in the Supplementary Methods. For studies which included patients of European and non-European ancestries, only data reported for non-Europeans were extracted. In instances where information provided in the published manuscript was unclear, we contacted the study authors by email for clarification but amongst the six emails sent out, no response was received, and therefore these 6 articles were excluded. If the exact number for a data item could not be extracted, meticulous estimation was undertaken where possible. All extracted data were presented and compared between T.H.C and J.E.Z, with any disagreements resolved by discussion to reach consensus.

### Data synthesis

Due to the heterogeneity of articles included in this systematic review and the small number of studies conducted in each ethnicity, it was impossible to perform a quantitative analysis, and so the findings are described in a narrative way and data extracted from each article presented in tables. No meta-analysis was undertaken.

### *In silico* prediction

*In silico* prediction was undertaken for all *DPYD* genetic variants evaluated in this systematic review and is described in the Supplementary Methods. The scoring thresholds and software weblinks of the *in silico* prediction tools used are summarised in Supplementary Table 3.

### Published *in silico* functional predictions and *in vitro* functional data

To acquire a more nuanced understanding of the *DPYD* variants identified in our systematic review, published data from previously developed *in silico* functional prediction models with high accuracy, the DPYD-varifier^53^ and the ADME-optimised Prediction Framework (APF) ^54,55^, were extracted (described in Supplementary Methods). In addition, functional data on DPD enzyme activity from *in vitro* experiments where HEK293T/c17, HEK293-Flp-In and 293FT cells were transiently expressed with *DPYD* variants and treated with either 5-FU or thymine were extracted ^42,45,56–59^.

## Results

### Identification and selection of articles

A detailed flow diagram showing the identification and selection process for study inclusion, according to the PRISMA statement, is depicted in Figure 1. All articles included were in English; none of the non-English articles met the criteria for inclusion.

**Figure 1:**
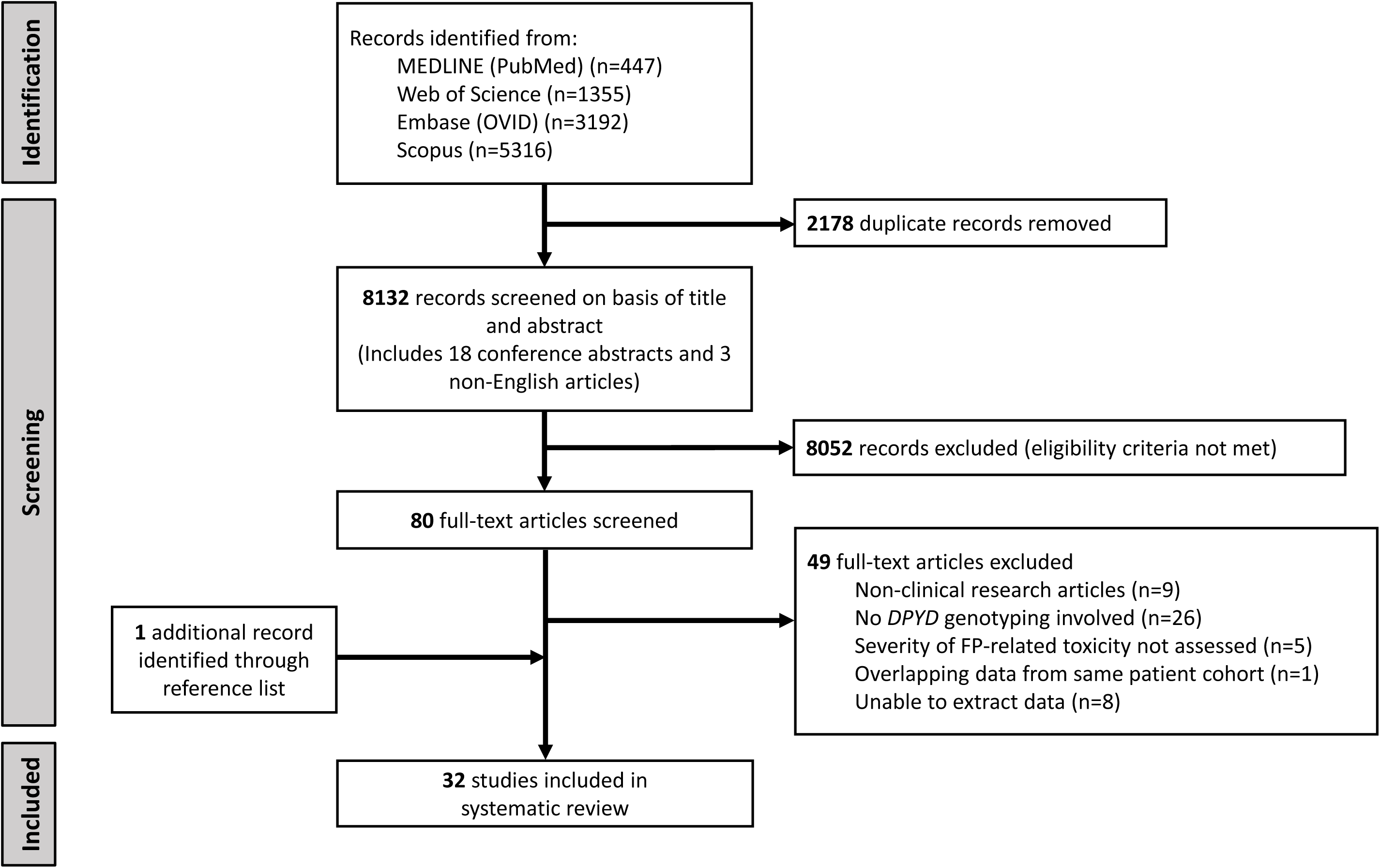
PRISMA flow diagram of study selection. Our search of four electronic databases identified a total of 10310 records, 447 from MEDLINE (PubMed), 1355 from Web of Science, 3192 from Embase (OVID), 5316 from Scopus. After removing 2178 duplicates, 8132 unique records remained which included 18 conference abstracts and 3 non-English articles. Following the title and abstract screening phase, 8052 records that did not meet the inclusion criteria were excluded. Full-text inspection of the remaining 80 articles identified 31 articles that met the eligibility criteria for inclusion. Screening the reference lists of these 31 articles identified one more relevant article, and so 32 articles were finally included in the present systematic review.

### Characteristics of included articles

Table 1 details the 32 included articles and a summary breakdown of the characteristics is provided in Supplementary Table 4. All articles were published between September 1998 and December 2022. Two studies were case series, 10 studies were case reports and 20 were cohort studies with an equal split between prospective and retrospective study designs. Patients were from 12 countries encompassing 5 ethnic groups: African American (United States), East Asian (China, Japan, Korea, Thailand), Latin American (Chile), Middle Eastern (Jordan, Lebanon, Saudi Arabia, Tunisia), and South Asian (Bangladesh, India, United States).

**Table 1.**
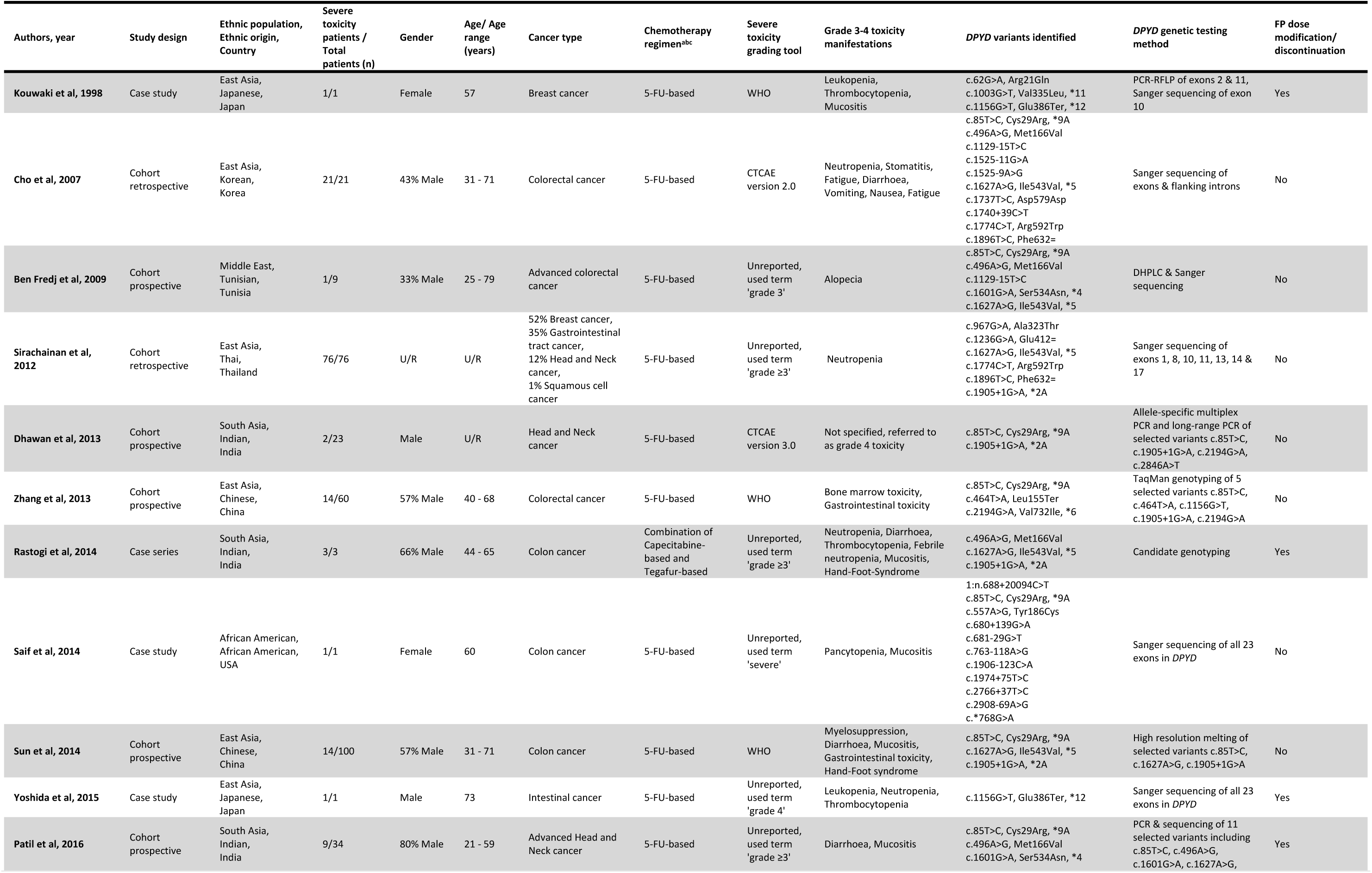
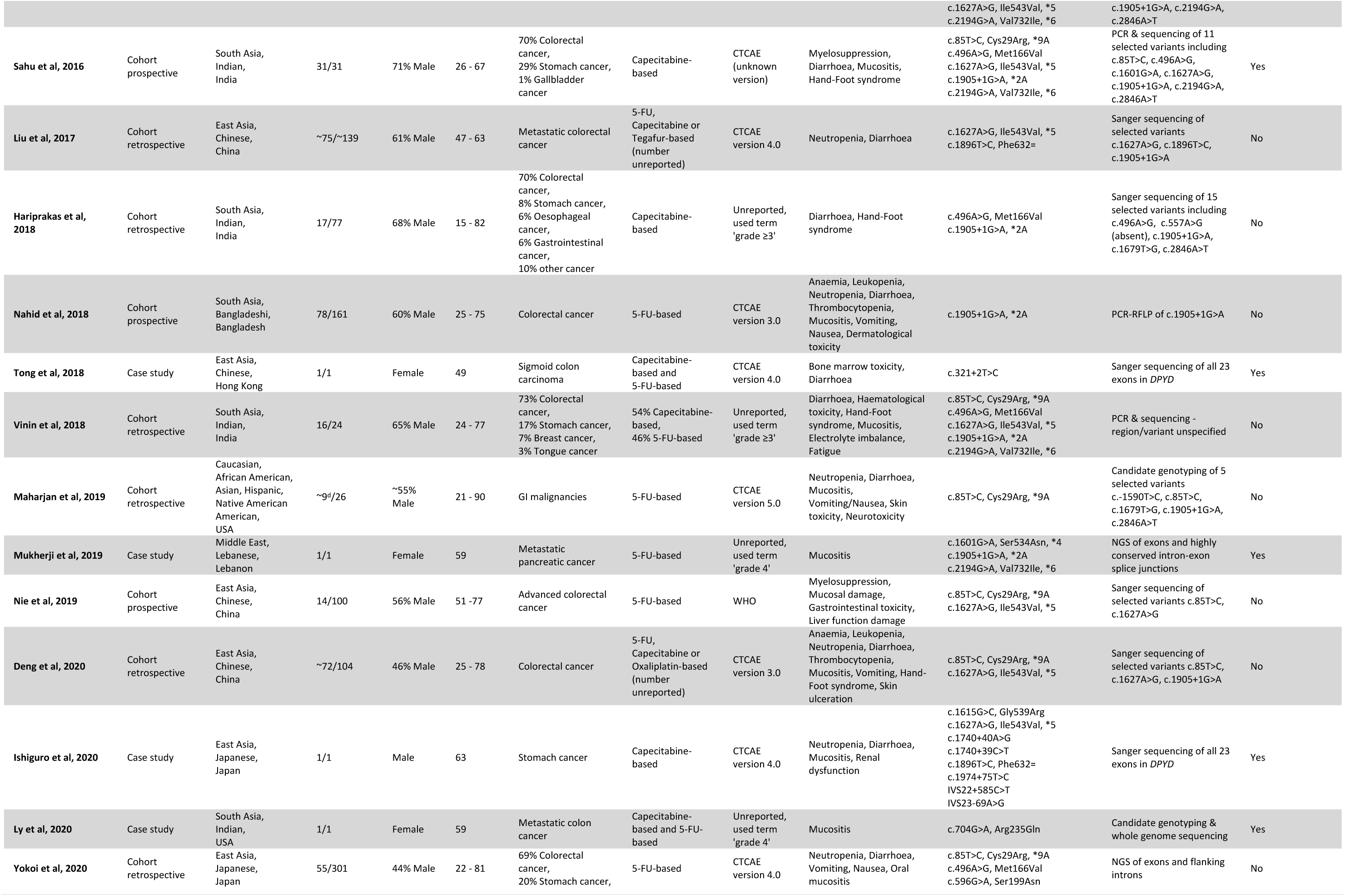
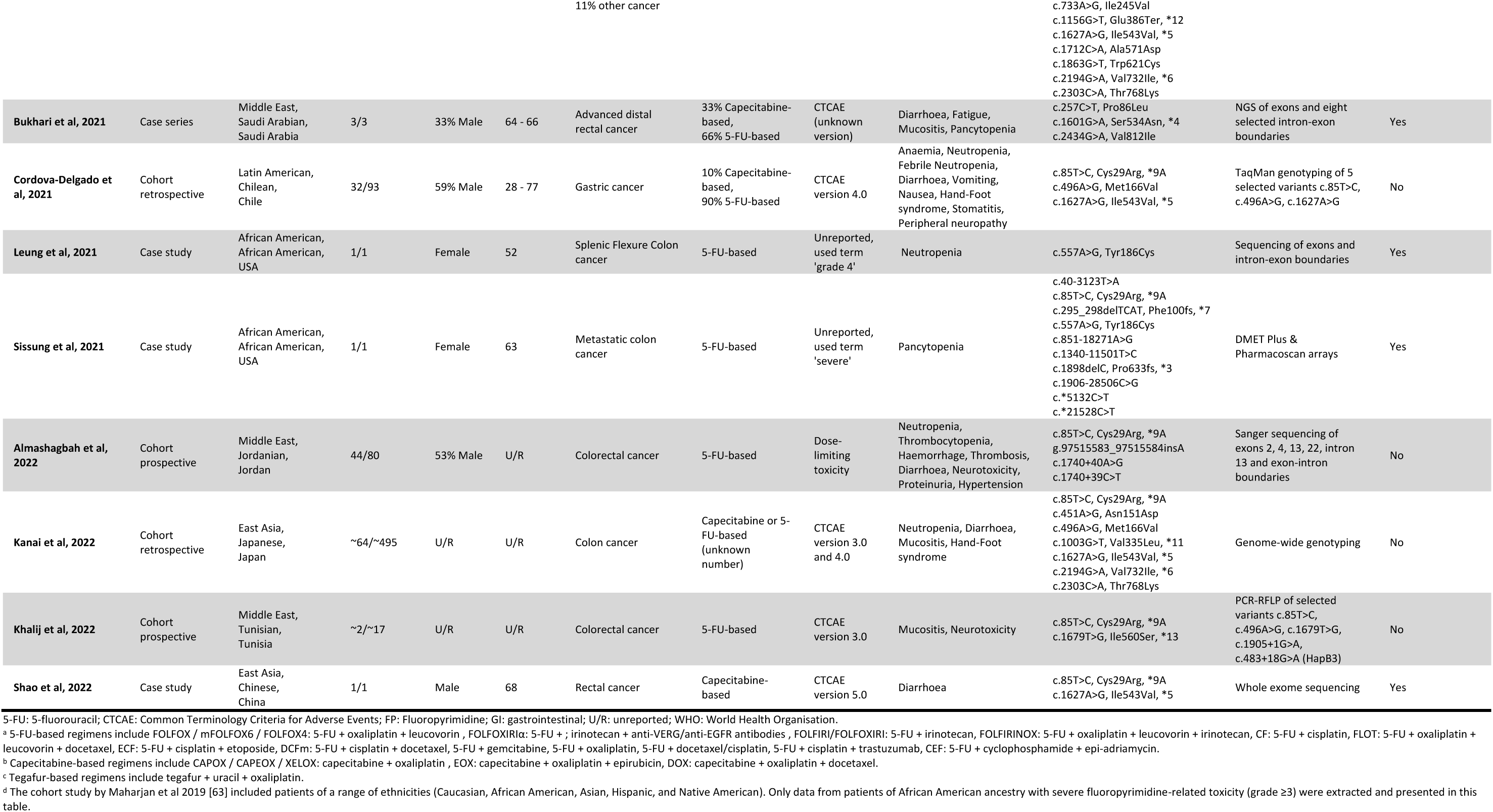
Characteristics of included studies.

Heterogeneity was present across the 32 articles included and are described in the Supplementary Results.

### Patient characteristics

A summary of the patient characteristics is presented in Table 2. A total of 1431 patients were included across the 32 studies. Their age ranged between 15 to 90 years, and slightly more men than women were enrolled in most studies. The most common type of tumour was colorectal cancer and most patients received either 5-FU or capecitabine-based combination chemotherapy treatment that included oxaliplatin. All patients were reported to have experienced grade 3 or higher fluoropyrimidine-related toxicities (as defined above). Clinical manifestations included haematological, gastrointestinal, dermatological, hepatic, neurological, and renal toxicities, with many with neutropenia, myelosuppression, diarrhoea, mucositis and hand-foot-syndrome.

**Table 2.**
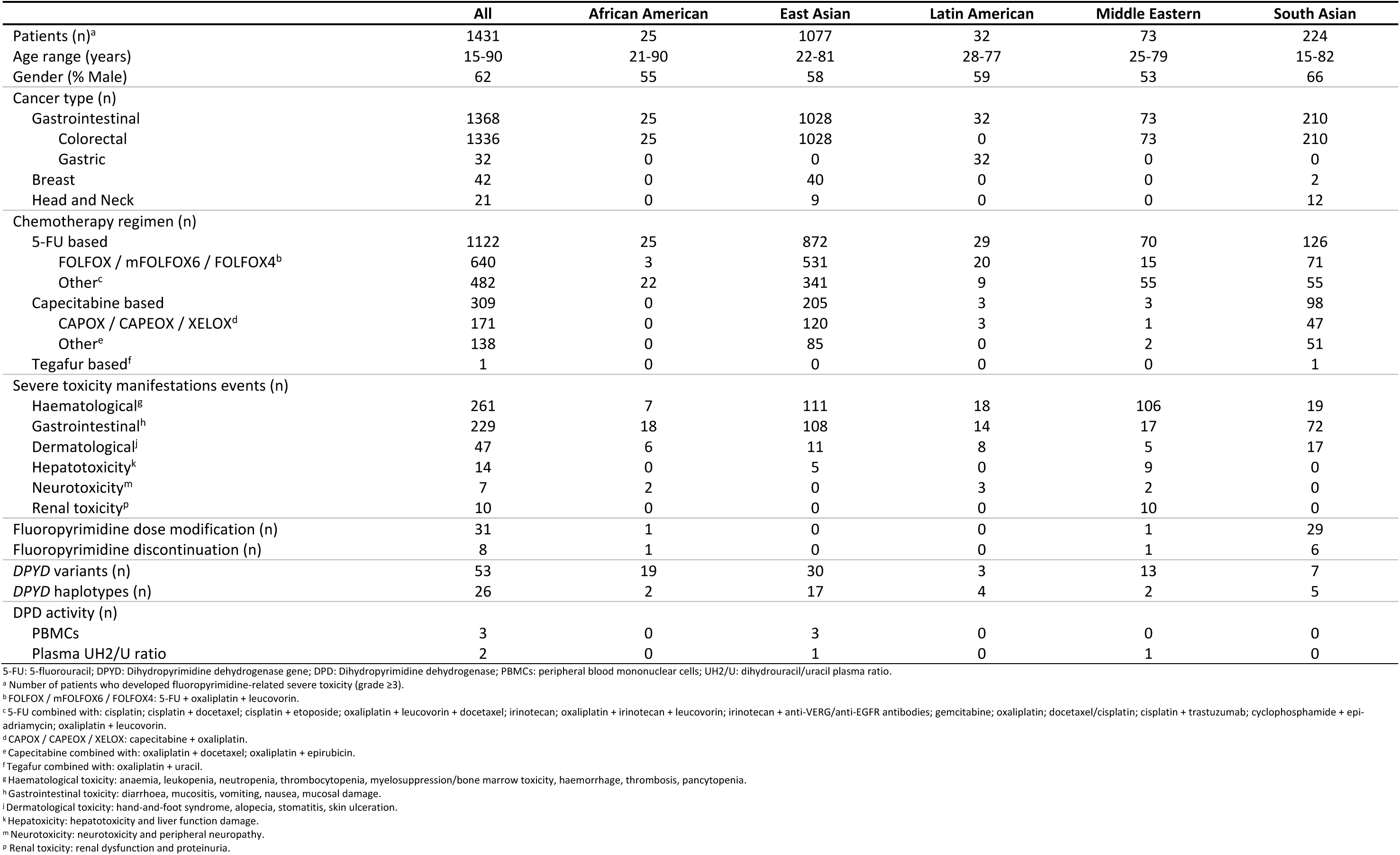
Patient characteristics.

### *DPYD* genetic variants, haplotypes and *in silico* predictions

Across the 32 included studies, a total of 53 *DPYD* genetic variants were reported, of which 20 have been reported in the CPIC guideline^6^ (Figure 2). Genotype counts of variants reported in patients with severe fluoropyrimidine-related toxicity across the 5 ethnicities with details of all extracted data items are presented in Supplementary Table 5. Our *in silico* prediction results for all 53 *DPYD* variants identified are summarised in Table 3 with scores obtained from each *in silico* prediction tool detailed in Supplementary Table 6. In addition, 13 studies reported a combination of *DPYD* genetic variants at individual patient-level and we were able to identify 26 haplotype combinations as presented in Supplementary Table 7. Subsequent paragraphs in this section will focus on variants which were reported in more than 1 individual in each ethnicity with either: (1) CPIC-reported decreased or loss of DPD enzyme function or (2) unreported DPD enzyme function in the CPIC guideline but predicted to be deleterious by >60% of the *in silico* tools we utilised. Variants which were excluded due to this filtering process and haplotype combinations are described in the Supplementary Results.

**Figure 2:**
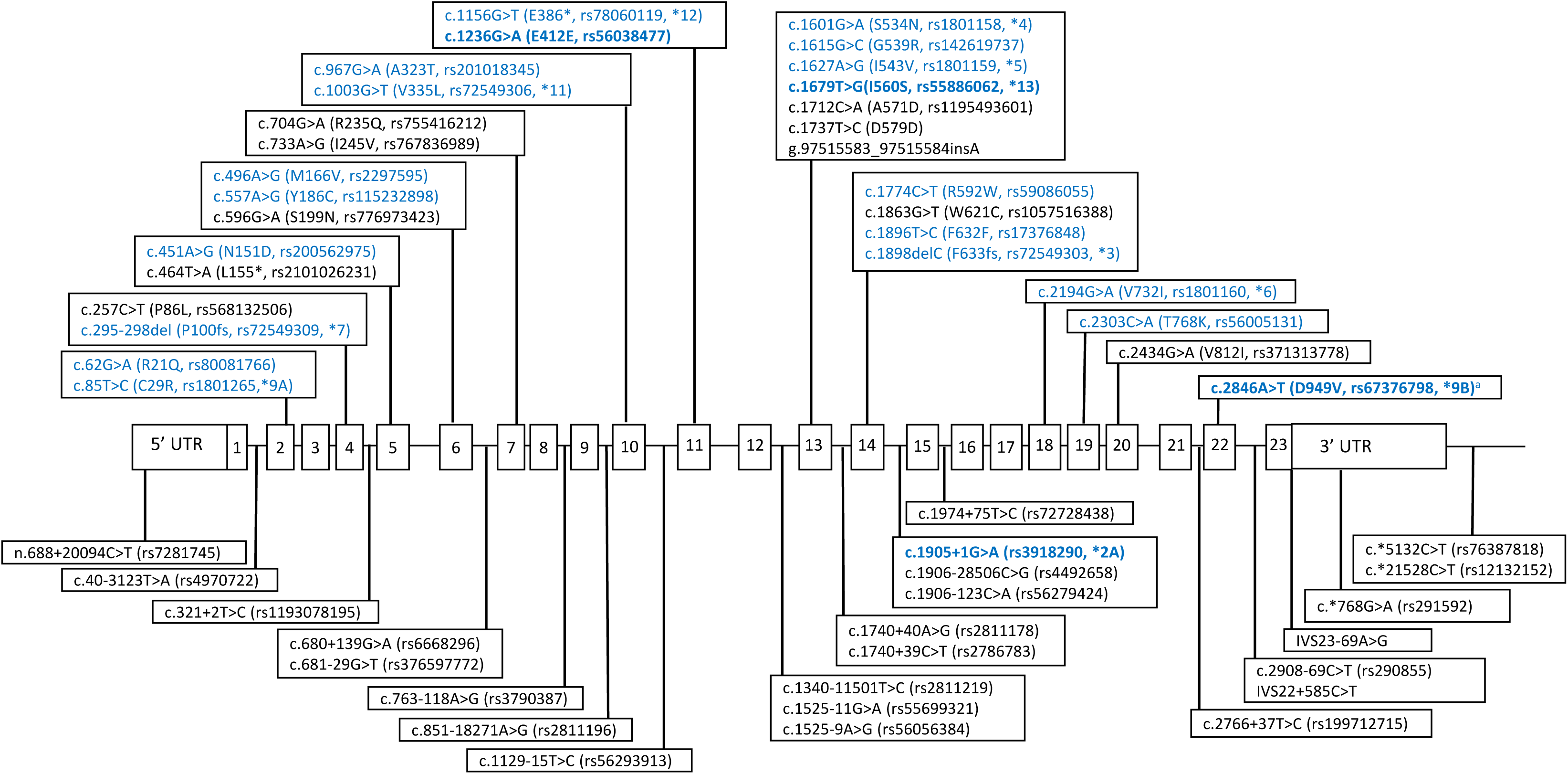
53 *DPYD* variants identified in our systematic review. Variants listed in the CPIC guideline are highlighted in blue. The four prominent European *DPYD* variants are in bold blue font. ^a^c.2846A>T was not identified in our systematic review.

**Table 3:**
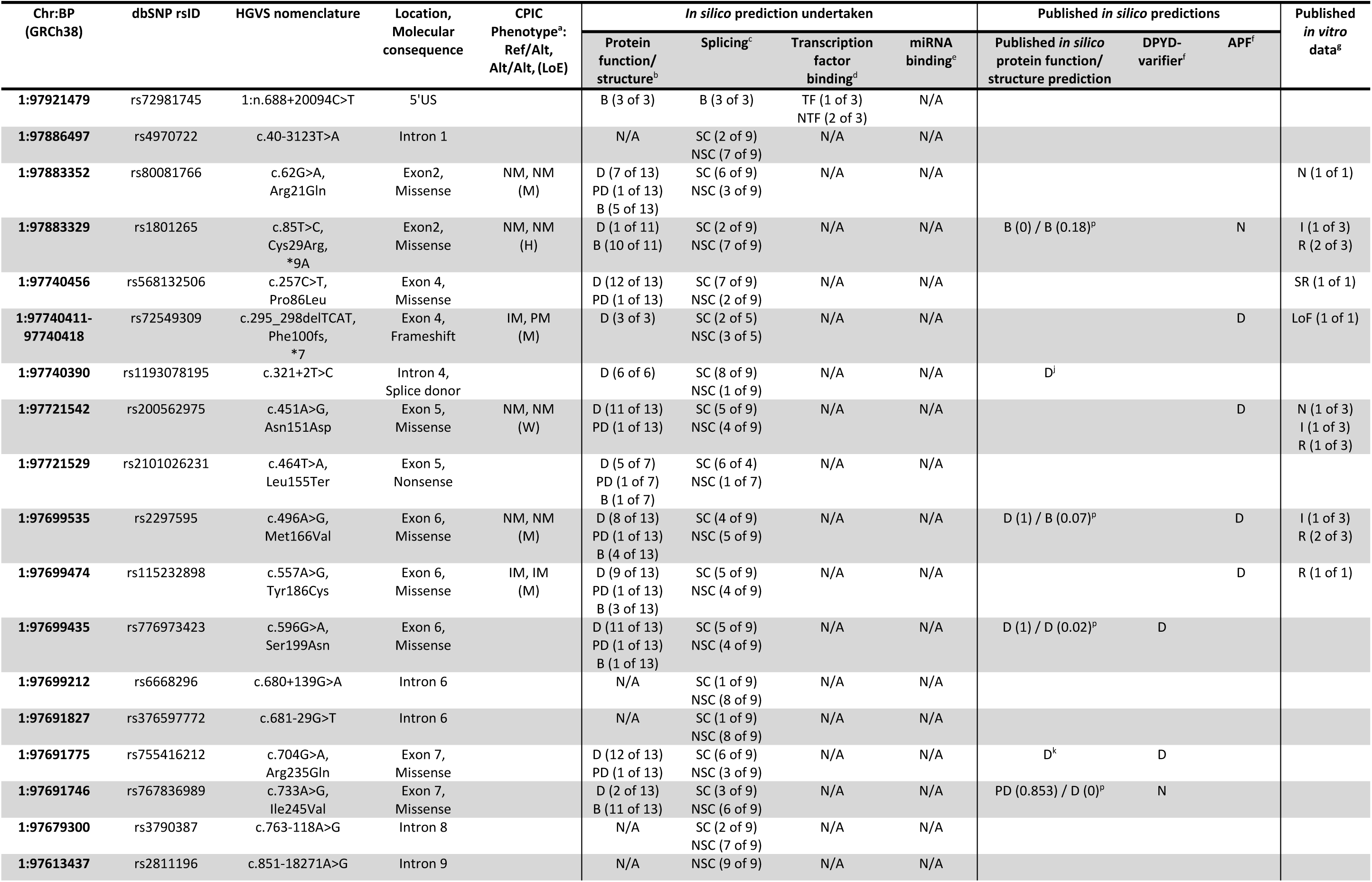
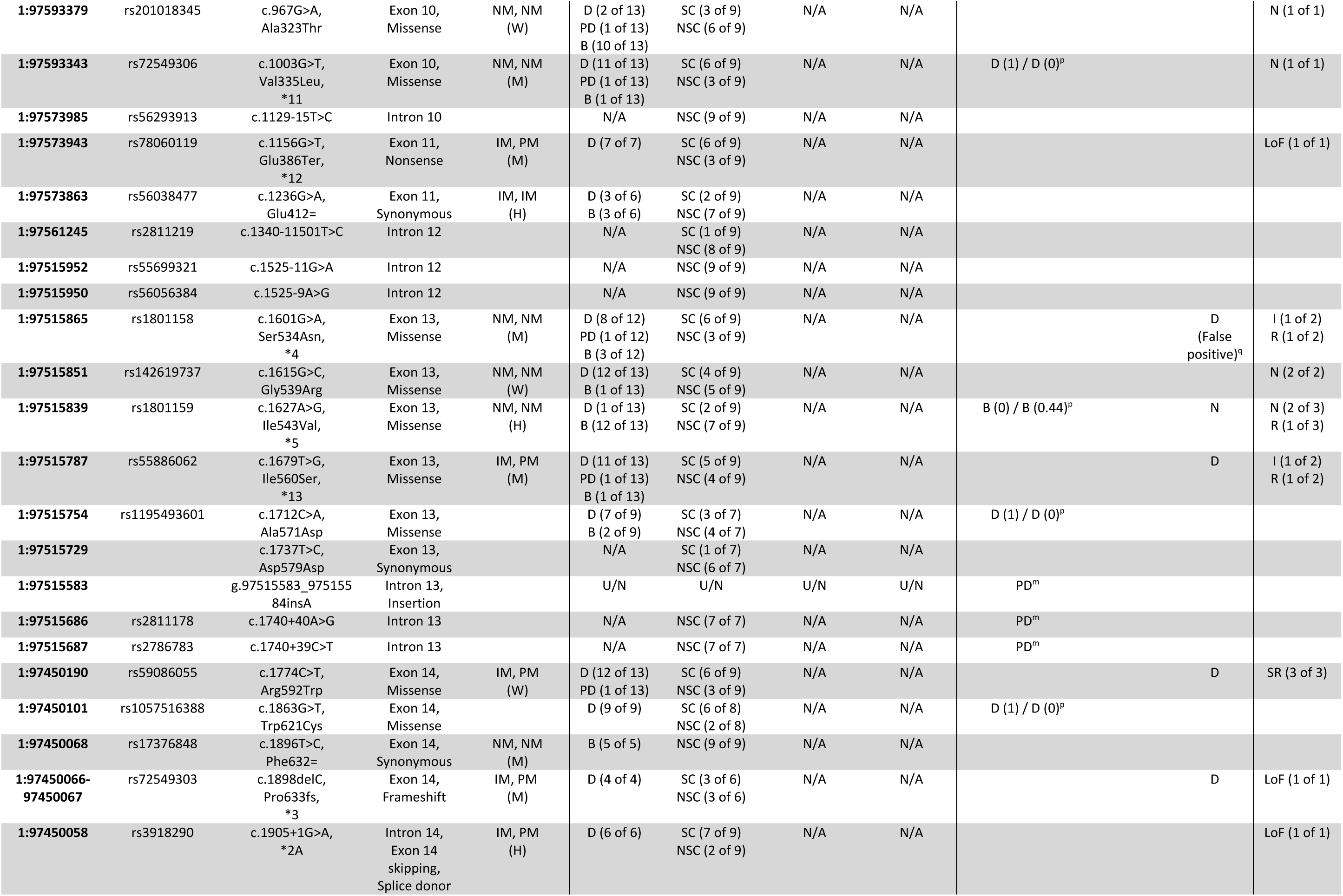
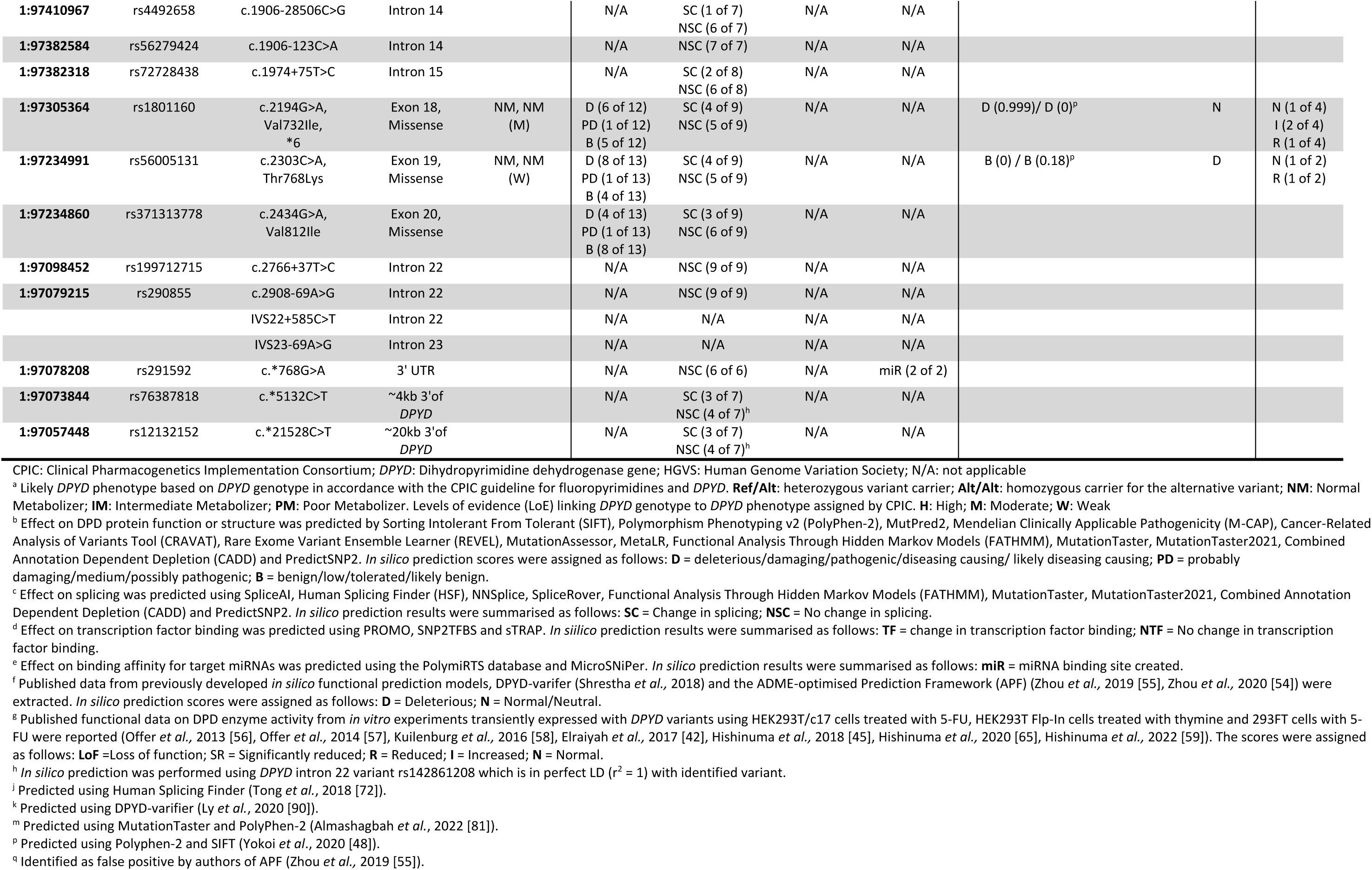
*In silico* predictions of *DPYD* variants evaluated in our systematic review.

#### African American

19 *DPYD* variants (2 missense, 2 frameshift, 11 intronic, one 5’-upstream, one 3’UTR, two 3’-downstream) were reported across 3 case studies ^60–62^ and 1 cohort study ^63^ conducted in patients of African American ancestry in the United States (Supplementary Table 5).

Heterozygous carriage of the missense variant c.557A>G (Tyr186Cys) was reported in all 3 case studies ^60,62,64^. This variant has a mean prevalence of ∼2% in reference populations of African descent (Supplementary Table 1) ^40,41^ and the presence of either 1 or 2 copies of the c.557A>G variant allele is considered to cause a decrease in DPD enzyme function (intermediate metaboliser) by the CPIC guideline with moderate strength of evidence. Up to 77% of the *in silico* prediction tools we utilised predicted this variant to be deleterious or probably damaging and this variant was classified as deleterious by APF (Table 3). *In vitro* functional analysis containing the Tyr186Cys amino acid substitution showed a ∼15% reduction in DPD enzyme activity relative to the wild-type (Table 3) ^57,65^. Maharjan and colleagues (2019) did not include c.557A>G genetic testing in their cohort of African American patients ^63^.

#### East Asian

A total of 30 *DPYD* variants (2 nonsense, 15 missense, 3 synonymous, 2 splice donor, and 8 intronic) were reported in patients of East Asian ancestry which included 5 cohort studies ^66–70^, 2 case reports from China ^71,72^, 2 cohort studies ^46,48^, 3 case reports from Japan ^73–75^, 1 cohort study from Korea ^76^, and 1 cohort study from Thailand ^77^ (Supplementary Table 5).

Amongst the 30 variants identified, 15 have been reported in the CPIC guideline including 3 loss of function variants, c.1156G>T (Glu386Ter), c.1774C>T (Arg592Trp) and c.1905+1G>A, with moderate, weak, and high strength of evidence respectively. Heterozygous carriers of 1 of these 3 variants lead to decreased enzyme function and are classified as intermediate metabolisers by CPIC; while homozygous carriers of either of these 3 variants lead to loss of enzyme function and are classified by CPIC as poor metabolisers. In reference populations of East Asian descent, these 3 variants are rare with zero MAF observed for c.1156G>T and c.1905+1G>A, and a MAF of 0.1% for c.1774C>T (Supplementary Table 1) ^40,41^.

Heterozygous carriage of the truncating c.1156G>T variant was reported in three Japanese patients, two from case reports who both exhibited >10 fold decrease in PBMC DPD enzyme activity in comparison to normal/healthy individuals ^74,75^, and one from a cohort study where heterozygous carriage of 1 of the 7 rare pathogenic *DPYD* variants, c.596G>A, c.733A>G, c.914C>A, c.1156G>T, c.1666A>C, c.1712C>A, or c.1863G>T was significantly associated with grade 3-4 toxicity in comparison to patients without the 7 rare variants (OR = unreported; *p* = 0.0271; Supplementary Table 5) ^48^. 100% of the *in silico* prediction tools we utilised predicted c.1156G>T to be deleterious and published *in vitro* expression analysis reported complete loss of DPD enzyme activity (Table 3) ^57,65^.

Two patients, one from a Korean cohort study and one from a Thai cohort study, were heterozygous for the nonsynonymous variant c.1774C>T ^76,77^. Up to 92% of the *in silico* prediction tools we utilised predicted c.1774C>T to be deleterious and the APF classified this variant as deleterious (Table 3). Previously published *in vitro* functional characterization of c.1774C>T reported a reduction in DPD catalytic activity compared to the wild-type (Table 3) ^45,57,59,65^.

Heterozygous carriers of the intron 14 splice donor variant c.1905+1G>A were reported in one Thai cohort patient ^77^ and 14 Chinese cohort patients in which significantly higher incidences of grade 3-4 myelosuppression, hand-foot syndrome, diarrhoea, gastrointestinal reactions and mucositis were observed (OR = unreported; *p* < 0.001 for each severe side effect) compared to wild-type carriers ^69^. 100% of the *in silico* prediction tools we utilised predicted this variant to be deleterious and published *in vitro* expression analysis reported c.1905+1G>A to be catalytically inactive (Table 3) ^56,65^.

Two Chinese patients from a cohort study, one with grade 4 bone marrow inhibition (BMI) and one with grade 4 BMI and grade 4 gastrointestinal toxicity, were reported to be heterozygous carriers for the nonsense variant, c.464T>A (Leu155Ter). This variant is not reported in the CPIC guideline. The DPD enzyme activity in PBMCs from both patients was ∼45% lower than that in non-carriers with Grade 1-2 toxicity (Supplementary Table 5) ^70^. In addition, when c.464T>A was analysed in composite with c.85T>C and c.2194G>A, the carriage of either c.464T>A, c.85T>C, and/or c.2194G>A was associated with an increased incidence of bone marrow toxicity (OR = 24; *p* = 0.0001) and gastrointestinal toxicity (OR = 8; *p* = 0.0019) in comparison to non-variant carriers (Supplementary Table 5) ^70^. 70% of the *in silico* prediction tools we used predicted the c.464T>A to be deleterious (Table 3, Supplementary Table 6). No allele frequency information in reference populations of East Asian descent and other ancestries has been reported for this variant (Supplementary Table 1) ^40,41^.

#### Latin American

Only 1 cohort study from Chile was identified in the Latin American population ^78^ and the authors examined 3 missense *DPYD* polymorphisms considered to have normal DPD enzyme function by the CPIC guideline, c.85T>C, c.496A>G and c.1627A>G (Supplementary Table 5, Supplementary Results).

#### Middle Eastern

13 *DPYD* variants (1 splice donor, 8 missense, 4 intronic) were reported in patients of Middle Eastern ancestry. There were 2 cohort studies from Tunisia ^79,80^, 1 cohort study from Jordan ^81^ 1 case report from Lebanon ^82^ and 1 case series from Saudi Arabia ^83^ (Supplementary Table 5). None of the variants passed our filtering process (Supplementary Results).

#### South Asian

7 *DPYD* variants (6 missense, 1 splice donor) were reported in patients of South Asian ancestry across 5 cohort studies from India ^84–88^, one Indian case series ^89^, one case study of an Indian patient in the USA ^90^, and 1 cohort study from Bangladesh ^91^ (Supplementary Table 5).

With a prevalence of 0.3-1.5% in reference populations of South Asian descent (Supplementary Table 1) ^40,41^, the splice donor variant c.1905+1G>A was reported in patients from Bangladesh and India ^84,85,87–89,91^. The Bangladeshi cohort study reported a significant association with anaemia (OR = 4.7, *p* = 0.042), neutropenia (OR = 6.47, *p* = 0.018), thrombocytopaenia (OR = 8.08, *p* = 0.05), nausea (OR = 10.06, *p* = 0.012), and diarrhoea (OR = 5.76, *p* = 0.026) when patients with grade 3-4 toxicities were compared to patients with grade ≤2 toxicities ^91^. The Bangladeshi cohort study genotyped for only the c.1905+1G>A variant, and whether there were other mutations was not investigated. One of the four Indian cohort studies reported a decreased incidence of mucositis (*p* = 0.016) and diarrhoea (*p* = 0.006) in *DPYD* variant carriers of either c.85T>C, c.496A>G, c.1627A>G, c.1905+1G>A and/or c.2194G>A after 50% capecitabine dose reduction in cycle 2 of chemotherapy ^87^.

## Discussion

This systematic review has identified numerous variants in the *DPYD* gene which have been reported in non-European individuals with severe toxicity associated with the use of fluoropyrimidines. In the UK and EU, testing for 4 *DPYD* genetic variants is undertaken before the use of fluoropyrimidines ^36,37^ – in England, we currently do 38,000 tests per year. This is an important success story for the implementation of pharmacogenomics, but there is still a need to improve the testing pathway, both in terms of increasing the number of genetic variants tested, and ensuring that we are not disadvantaging particular ethnic groups.

It is interesting to note that our systematic review has identified 3 of the 4 *DPYD* variants tested in the UK and EU^36,37^, in non-European individuals. The c.1905+1G>A variant, which leads to exon 14 skipping, has been reported in 1 Thai ^77^, 14 Chinese ^69^, 1 Lebanese ^82^, 7 Bangladeshi ^91^ and 18 Indian ^84–89^ patients with fluoropyrimidine-related toxicity. The frequency of this variant is 0% in East Asian reference populations, 0.3% in Middle Eastern reference populations, and 0.3-1.5% in South Asian reference populations ^40,41^. The c.1679T>G and c.1236G>A/HapB3 variants have been reported in 1 Tunisian patient ^80^ and 1 Thai patient ^77^, respectively. The prevalence of c.1679T>G is 0% in Middle Eastern reference populations ^41^ and the frequency of c.1236G>A/HapB3 ranges from 0.01-0.1% in East Asian reference populations ^41^. According to the 2021 UK census ^92^, South Asians, East Asians, and Arabs represent 6.7%, 1.3%, and 0.6% of the UK population, respectively, and thus they will benefit from the genetic testing which is offered to all patients in the UK if they require treatment with 5-FU or its analogues.

Clearly, there are other variants in these ethnic groups which need further investigation. For example, in South Asians and Middle Easterners, our systematic review identified single occurrence of missense variants c.704G>A (p.Arg235Gln, rs755416212) ^90^ and c.257C>T (p.Pro86Leu, rs568132506) ^83^, respectively. These variants are not reported in the CPIC guideline but are predicted to be deleterious by >80% of the *in silico* tools we used, with one research study reporting significant reduction of DPD activity *in vitro* with the c.257C>T variant ^42^. Further functional work and greater interrogation of patients who have had toxicity is warranted to confirm these findings and to identify other functionally relevant variants.

It is important to briefly consider some other variants. First, c.464T>A (p.Leu155Ter, rs2101026231), a nonsense variant, located on exon 5, causes the replacement of leucine 155 by a stopping codon, resulting in a truncated protein. This variant was first reported in a Spanish patient who died from severe, multi-system toxicity following the first administration of 5-FU for adjuvant colon cancer therapy ^93^. Two Chinese patients with 5-FU-related severe toxicity who both patients exhibited ∼45% lower DPD enzyme activity in PBMCs compared to non-carriers have also been reported ^70^. This variant is not included in the current CPIC guideline and its allele frequency across global ethnic populations in the 1000 Genomes and gnomAD databases is currently unknown, but our *in silico* analysis predicted this variant to be deleterious or probably damaging but further *in vitro* functional work is required to confirm the impact of this variant on DPD enzyme activity. Second, the exon 11 truncating variant, c.1156G>T (p.Glu386Ter, rs78060119, *12), leads to premature protein truncation at amino acid position 386 and is classified as a loss-of-function variant with moderate evidence level by the CPIC guideline. Heterozygous carriage of this variant was detected in three Japanese patients with severe fluoropyrimidine-related toxicity across 2 case studies and 1 cohort study identified in our systematic review ^48,74,75^. *In vivo* and *in vitro* studies of this variant observed over 90% reduction in DPD activity ^57,74,75^. Currently, there is no guideline or mandate for *DPYD* testing before fluoropyrimidine treatment in Japan but *DPYD* genetic testing that includes the c.1156G>T variant is available in several hospital pharmacies and can be requested by the attending physician. Data from the latest gnomAD release (v4.0) showed very low prevalence (<0.005%) of this variant across East Asian, Admixed American, South Asian, and European populations. Third, the c.1774C>T (p.Arg592Trp, rs59086055) variant has a prevalence of 0.1% in East Asian reference populations ^40,41^; heterozygous carriage of c.1774C>T was detected in 1 Korean patient ^76^ and 1 Thai patient ^77^ in our systematic review. This exon 14 missense variant causing Arg592Trp substitution is considered a loss-of-function variant by the CPIC guideline with weak evidence. No *in vivo* data have been published for this variant but *in vitro* functional work reported over 90% reduction in DPD activity ^45,57,59,65^. These variants seem to be important but further work is required both to understand the functional relevance of these variants, and identify other variants in East Asian individuals, to improve the prediction of fluoropyrimidine-related toxicity in the different ethnic groups that constitute East Asian populations in the UK and globally.

Our systematic review has identified 3 case studies detecting the c.557A>G variant (rs115232898, p.Tyr186Cys) in African Americans with severe 5-FU-related toxicity ^60–62^, one of which was fatal ^61^. In addition, in an editorial which was not eligible for inclusion in our systematic review, this variant was reported in an African-Caribbean patient with severe 5-FU-related toxicity ^94^. This is a nonsynonymous variant located on exon 6 where carriers have 46% lower DPD enzyme activity in PBMCs than non-carriers ^95^. *In vitro* functional analysis of DPD containing the Tyr186Cys amino acid substitution has shown a ∼15% reduction in DPD activity relative to wildtype ^57^. Data from the 1000 Genomes Project Phase 3 confirms that c.557A>G is mainly found in African populations (Afro-Caribbeans in Barbados, African Americans in southwest United States, Yoruba in Ibadan (Nigeria), Luhya in Webuye (Kenya), Gambian in Western Divisions in the Gambia, Mende in Sierra Leone, and Esan in Nigeria), with allele frequency ranging between 1-4% ^40^. This variant is virtually non-existent in Europeans, East Asians and South Asians. In the United States, the Mayo Clinic and several commercial laboratories includes c.557A>G in their pre-treatment *DPYD* testing to identify individuals at increased risk of toxicity when considering fluoropyrimidine chemotherapy treatment. However, this variant is currently not included in the UK NHS *DPYD* genetic testing. In the 2021 UK Census, 4% (2.4 million) of the total population in England and Wales identified their ethnic group within the "Black, Black British, Black Welsh, Caribbean or African" category ^92^.

Our systematic review also shows that few novel variants in the *DPYD* gene have been reported in Middle Eastern ^81^ populations with a paucity of data in Latin American populations ^78^, highlighting the need for more studies in these populations. Indeed, further studies are needed in all populations (European and non-European) to fully understand the spectrum of harmful mutations which occur in this gene. This will require careful identification and assessment of patients with toxicity caused by 5-FU or its analogues, and subsequent sequencing of the *DPYD* gene together with functional characterisation of any mutations identified. To this end, we have initiated a programme of work (called “DPYD-International”) which has the aim to identify affected patients globally so that evidence can be generated to optimise the pathway for *DPYD* genetic screening to maximise benefits for all populations and minimise any unintended inequalities.

Previous studies have shown that *DPYD* intermediate and poor metabolizers receiving conventional doses of fluoropyrimidine are at significantly higher risk for severe toxicity and treatment-related mortality ^31,32^ and pre-treatment testing followed by genotype-guided dose reduction in variant carriers significantly reduces toxicity and mortality risks ^31–35^, and associated hospitalisations ^32,96–98^. This strategy has also been shown to be cost-effective. For example, a UK-based study of an extended *DPYD* genetic panel showed that genotyping was dominant over standard of care, with a saving of £78,000 per patient over a lifetime ^99^. Two other studies, one from Canada^100^ and another from Iran^101^, have also shown pre-prescription *DPYD* genotyping to be cost saving, while studies from the US^96^ and Spain^102^ showed it to be cost-effective.

Our systematic review has limitations. We had to rely on observational studies, including case reports, to identify affected patients. Clearly this represents selective reporting, and many patients with important variants are either not reported, or more likely not genotyped or sequenced. It is therefore important to identify and sequence these patients to evaluate the full spectrum of mutations associated with toxicity from 5-FU or its analogues. For many of the variants identified, the functional consequences are unknown. In this review, we have undertaken a comprehensive *in silico* evaluation of the likely functional consequences of the mutations, but further functional evaluation will be needed for many of the variants. Notably, our systematic review has identified a number of patients carrying more than one *DPYD* variant and in particular one African-American carrying 2 loss-of-function variants c.295_298delTCAT and c.1898delC in addition to the decreased function variant c.557A>G (Supplementary Results)^62^; how the co-expression of functional *DPYD* variants affects overall DPD activity and the consequences for the severity of fluoropyrimidine-related toxicity remains to be elucidated. Our focus has been on the *DPYD* gene, but there are other potential genes (e.g. MIR27A, TYMS, ENOSF1, MHTFR) which may be important in predisposing to toxicity from the fluoropyrimidines, and these will need a separate evaluation.

In conclusion, our systematic review has focused on non-European patients and has identified numerous variants in the *DPYD* gene which have been reported in patients with severe toxicity after treatment with 5-FU or its oral analogues. The UK is an increasingly multi-cultural and ethnically diverse society but we test for 4 variants which have been identified from studies undertaken in European populations. However, our analysis shows that 3 of these 4 variants are also important in South Asian, East Asian and Middle Eastern individuals. From the evidence gathered, and based on practice elsewhere in the world, we feel that it would be important to extend *DPYD* genetic testing in the UK NHS to include the c.557A>G variant which has been identified in individuals of African ancestry. The other variants described in this systematic review need further evaluation for incorporation into the testing pathways either in the UK or elsewhere, but of course, if sequencing becomes the standard method for characterising *DPYD* variation, we hope the information contained within this systematic review will be of use to diagnostic labs and policy makers.

## Additional Information

### Authors’ contributions

Conceptualization, M.P. and E.J.Z.; Methodology, T.H.C. and E.J.Z.; Data review, T.H.C. and E.J.Z; *In silico* analysis, T.H.C.; Writing – Original Draft Preparation, T.H.C. and E.J.Z.; Writing – Review and Editing, T.H.C., E.J.Z. and M.P.; Supervision, E.J.Z. and M.P. All authors have read and agreed to the published version of the manuscript.

### Ethics approval and consent to participate

Ethics approval was not required for this review.

### Data availability

Data used in this review is provided in Supplementary Appendices; any additional data are available upon request to the corresponding author.

